# Artificial intelligence-enhanced Electrocardiography Score for Perioperative Risk Assessment in Non-cardiac Surgery

**DOI:** 10.1101/2025.05.20.25328021

**Authors:** Hong-Mi Choi, Yerin Kim, Joonghee Kim, Jiesuck Park, Ji Hyun Lee, Yeonyee E. Yoon, Il-Young Oh, In-Ae Song, Youngjin Cho

## Abstract

**Background:** The role of electrocardiography (ECG) has been limited in the preoperative risk evaluation in noncardiac surgery due to its low prognostic value. Recent advances in artificial intelligence (AI) have enabled the extraction of subtle features from ECG that can be used in risk prediction.

**Objectives:** This study aimed to evaluate the utility of an AI-enabled ECG (QCG-Critical score) in predicting 30-day postoperative mortality in non-cardiac surgery and compare its performance with traditional perioperative risk-assessment tools.

**Methods:** A retrospective cohort of 46,135 non-cardiac surgeries at a tertiary center between 2020 and 2021 was analysed. Preoperative ECG images acquired within 30 days before surgery were input to a previously developed CNN-based deep-learning algorithm to generate QCG-Critical score, and its ability to predict 30-day in-hospital mortality was evaluated. Secondary outcomes included 7-day mortality, prolonged mechanical ventilation, unplanned percutaneous coronary intervention, and heart failure within 30 days postoperatively.

**Results:** The 30-day postoperative mortality rate was 0.34%. The QCG-Critical score model showed strong predictive performance (AUROC: 0.909) predicting postoperative mortality, outperforming the ESC guidelines (0.728) and RCRI (0.725) and performing comparably to the ASA classification (0.886). The QCG-Critical score model remained predictive across clinical subgroups and demonstrated superior performance in predicting secondary outcomes: 7-day mortality, unplanned percutaneous coronary intervention, mechanical ventilation, and heart failure.

**Conclusion:** The preoperative QCG-Critical score accurately predicted postoperative mortality and other adverse outcomes, outperforming conventional risk stratification tools.

The QCG-Critical score may serve as a fast, accessible, and integrable tool for perioperative risk assessments in routine surgical care.

**Condensed Abstract:** The AI-driven QCG-Critical score predicted 30-day postoperative mortality after non-cardiac surgery with excellent accuracy (AUROC 0.909), outperforming traditional risk models. It also showed strong predictive power for adverse outcomes including early mortality, unplanned PCI, and heart failure. Unlike existing models, the QCG-Critical score requires only ECG images and no additional clinical inputs, enabling rapid, objective, and scalable perioperative risk assessment. This AI-enabled approach may optimize surgical decision- making and resource utilization.

## INTRODUCTION

More than 200 million noncardiac surgeries are performed annually worldwide.^1^ To minimize unexpected postoperative cardiac events, several clinical risk-stratification tools have been developed and widely utilized.^2^ Despite the increasing complexity of these tools with the incorporation of additional variables to improve predictive accuracy, their overall performance remains suboptimal.^2,3^ Owing to advances in surgical techniques and increased life expectancy, an increasing number of older patients with multiple comorbidities are undergoing surgery. Simultaneously, the mortality rate of non-cardiac surgery is lower than the rate from several decades ago.^4,5^ Thus, this demographic shift may limit the utility of conventional risk-stratification strategies that were originally developed for different patient populations and clinical contexts.

Electrocardiography (ECG) is a simple, non-invasive assessment tool that has long been used for preoperative cardiac evaluation. Despite its low cost, accessibility, and widespread availability, owing to its limited prognostic value, current guidelines do not recommend routine ECG-based screening preoperatively for low-risk surgeries or in asymptomatic patients without risk factors.^2,3^ This limited utility may have originated from the narrow scope of information that is usually derived from ECG, such as rhythm disturbances or evidence of acute myocardial infarction, which may not sufficiently reflect the patient’s overall perioperative risk. Consequently, the role of the ECG in preoperative risk assessment remains unclear.^6,7^

Recent advancements in artificial intelligence (AI) and machine learning (ML) have enabled novel applications of ECG beyond that of conventional interpretations. AI-enhanced ECG models can extract subtle and often imperceptible features from standard 12-lead ECGs and thereby enable predictions beyond those that clinicians can visually detect. A growing body of literature has demonstrated the utility of AI-enabled ECGs for predicting various patient characteristics, structural abnormalities, and future cardiovascular outcomes with considerable accuracy.^8^

In this study, we evaluated the utility of AI-derived ECG parameters (QCG) in predicting perioperative 30-day mortality in patients undergoing non-cardiac surgery. Furthermore, we compared the predictive performance of the QCG-based model with that of existing perioperative risk-prediction tools.

## METHODS

### Study population

This single-centre, retrospective cohort study included patients who underwent elective non-cardiac surgery at Seoul National University Bundang Hospital between 2020 and 2021. Patients who underwent ECG within the 30 days before the surgery were included in this study. The exclusion criteria were as follows: 1) absence of preoperative ECG within the 30 days preceding the surgery, 2) elective cardiac surgery or extracorporeal membrane oxygenation-related surgery, 3) simple surgical procedures, such as nerve block, 4) surgeries undertaken under local anaesthesia, 5) surgery for a deceased organ donor, 6) rigid bronchoscopy, 7) dental surgery, and 8) multiple surgeries during the index admission. After applying these criteria, a total of 46,135 surgeries in 41,218 patients, which represents the total number of ECG–surgery pairs, was analysed in this study (**Supplementary** Figure 1).

### Data collection

All ECG recordings used consistent settings (25 mm/s speed and 10 mm/mV voltage gain), and the ECG image files in JPEG format were processed for QCG score extraction. Clinical data, including surgery-related information, anaesthesia records, laboratory results, and outcomes, were collected by reviewing electronic medical records. For comparative analysis, the surgical risk classification from European Society of Cardiology (ESC) guideline,^2^ the Revised Cardiac Risk Index (RCRI),^9^ and the American Society of Anesthesiologists Physical Status Class (ASA)^10^ were applied based on the type of surgery and history (Supplementary Table S1). This observational study follows the STROBE reporting guideline,^11^ and this study adheres to the TRIPOD reporting guideline.^12^

The study protocol was approved by the Institutional Review Board of Seoul National University Bundang Hospital (IRB No. B-2409-927-106), which waived the requirement for written informed consent owing to the retrospective nature of the study. All the clinical investigations were conducted in accordance with the principles of the Declaration of Helsinki.

### Endpoints

The primary outcome was in-hospital mortality within 30 days after the index surgery (30- day mortality rate). The secondary outcomes were in-hospital mortality within 7 days of index surgery (7-day mortality), unplanned percutaneous coronary intervention (PCI) within 30 days postoperatively, elevated troponin I within the postoperative 30-day period, mechanical ventilation for ≥3 days within 30 days postoperatively, and a possible acute decompensated heart-failure event, defined as either an N-terminal pro B-type natriuretic peptide level ≥300 pg/mL or the use of intravenous furosemide, within 30 days postoperatively after the index surgery.

### AI algorithm

A deep learning-based AI-ECG analysis system, a Quantitative ECG (QCG^TM^) analyser, was utilized to predict postoperative outcomes in this study. The QCG system is an AI analyser that uses 2D-ECG images (in JPG, PNG, or PDF formats) as input data and outputs the probability of specific downstream tasks as numerical values, ranging from 1 to 100. Using a modified convolutional neural network with residual connections, squeeze excitation modules, and a nonlocal block, the models were pretrained by using self-supervised learning schemes and were fine-tuned using multitask learning schemes on 47,194 annotated ECG images, from Seoul National University Bundang Hospital, that were recorded between 2017 and 2019. The tasks included the classification of rhythms and the production of 10 digital biomarkers (QCG scores) that correlated with various medical emergencies. This set of the QCG scores is bundled together and provided through an application named ‘ECG buddy,’ which was approved by the Korean Ministry of Food and Drug Safety (January 2024).

Detailed explanations of the AI algorithms and validation studies have been previously published.^13–16^ Among the 10 QCG scores, the QCG-Critical score, which was originally developed to identify patients at risk of critical illnesses, such as shock, respiratory failure, or cardiac arrest, was used to predict postoperative outcomes in this study.

### Statistical analysis

Continuous variables are presented as means with standard deviations or medians with interquartile range (IQR), as appropriate. Categorical variables are presented as frequencies with percentages. Intergroup comparisons were performed using the independent two-sample *t*-test for continuous variables and the chi-square test for categorical variables. When assumptions for parametric tests were not met, the Mann–Whitney *U* test was applied for continuous variables, and Fisher’s exact test was used for categorical variables with small sample sizes.

Given the presence of duplicate patients who underwent multiple surgeries during the enrolment period, we employed generalized estimating equation (GEE) models to develop a prediction model. Complete-case analysis was performed, excluding individuals with missing data in any variable included in the multivariate model. Moreover, multivariate GEE models were constructed to estimate the additive predictive power of various clinical variables, and the results were expressed as odds ratios (ORs) with 95% confidence intervals (CI).

The primary measure of model performance for predicting outcomes was the area under the receiver operating characteristic curve (AUROC). AUROC comparisons between different risk stratification tools and the QCG were conducted using DeLong’s tests. For sensitivity analysis, we repeated the same analysis on a subset of participants that included only one surgery per individual. For the subgroup analyses, interaction terms were introduced to assess the consistency of QCG performance across different patient categories.

Statistical significance was defined as a two-sided *p*-value <0.05. All statistical analyses were performed using the R software, version 4.4.1 (https://www.R-project.org).

## RESULTS

### Baseline characteristics

Baseline characteristics of the clinical and surgery-specific variables in the study population are presented in Table 1. The mean (SD) age was 56.6 (16.1) years, and 44.6% of the participants in the cohort were male. There were 1773 (3.8%) and 1515 (3.3%) patients with a history of ischemic heart disease and cerebrovascular events, respectively.

**Table 1.** Baseline characteristics of patients.

|  | Overall (N=46,135) |
| --- | --- |
| Age | 56.56 (16.14) |
| Male Sex | 20561 (44.6) |
| Previous history |  |
| Diabetes mellitus | 5904 (12.8) |
| Hypertension | 10759 (23.3) |
| Heart failure | 410 (0.9) |
| Ischemic heart disease | 1772 (3.8) |
| Cerebrovascular event | 1525 (3.3) |
| Smoking |  |
| Nonsmoker | 28436 (61.6) |
| Smoker | 9904 (21.5) |
| Unknown | 7795 (16.9) |
| Laboratory findings |  |
| Hemoglobin (g/dL) | 13.26 (1.86) |
| WBC count ($\times 10^3/\mu\text{L}$) | 6.45 (5.33–7.86) |
| Platelet count ($\times 10^3/\mu\text{L}$) | 241 (201.00–286.00) |
| BUN (mg/dL) | 15 (12.00–18.00) |
| Creatinine (mg/dL) | 0.74 (0.62–0.91) |
| AST (U/L) | 25 (20.00–31.00) |
| ALT (U/L) | 20 (14.00–30.00) |
| Albumin (g/dL) | 4.3 (4.10–4.50) |
| PT INR | 0.96 (0.93–1.01) |

Abbreviations: WBC, white blood cell; BUN, blood urea nitrogen; AST, aspartate aminotransferase; ALT, Alanine transaminase; PT INR, prothrombin time/international normalized ratio.

The surgery-related factors are listed in Table 2. Overall, 33,386 (72.8%) surgeries were performed under general anaesthesia. The mean (SD) ASA classification was 1.90 (0.73); moreover, 8,191 (18.8%) patients had an ASA classification ≥3. A total of 1,929 (4.2%) patients were classified as those with a high risk based on an RCRI ≥2, which represents more than 10% of major cardiac event risk. High-risk surgery, classified according to the ESC guideline, accounted for 10.7% of all surgeries. Emergency surgery was performed in 2,802 cases (6.1%). The composition of the surgical departments that were included in this study is shown in Supplementary Figure S2.

**Table 2.**
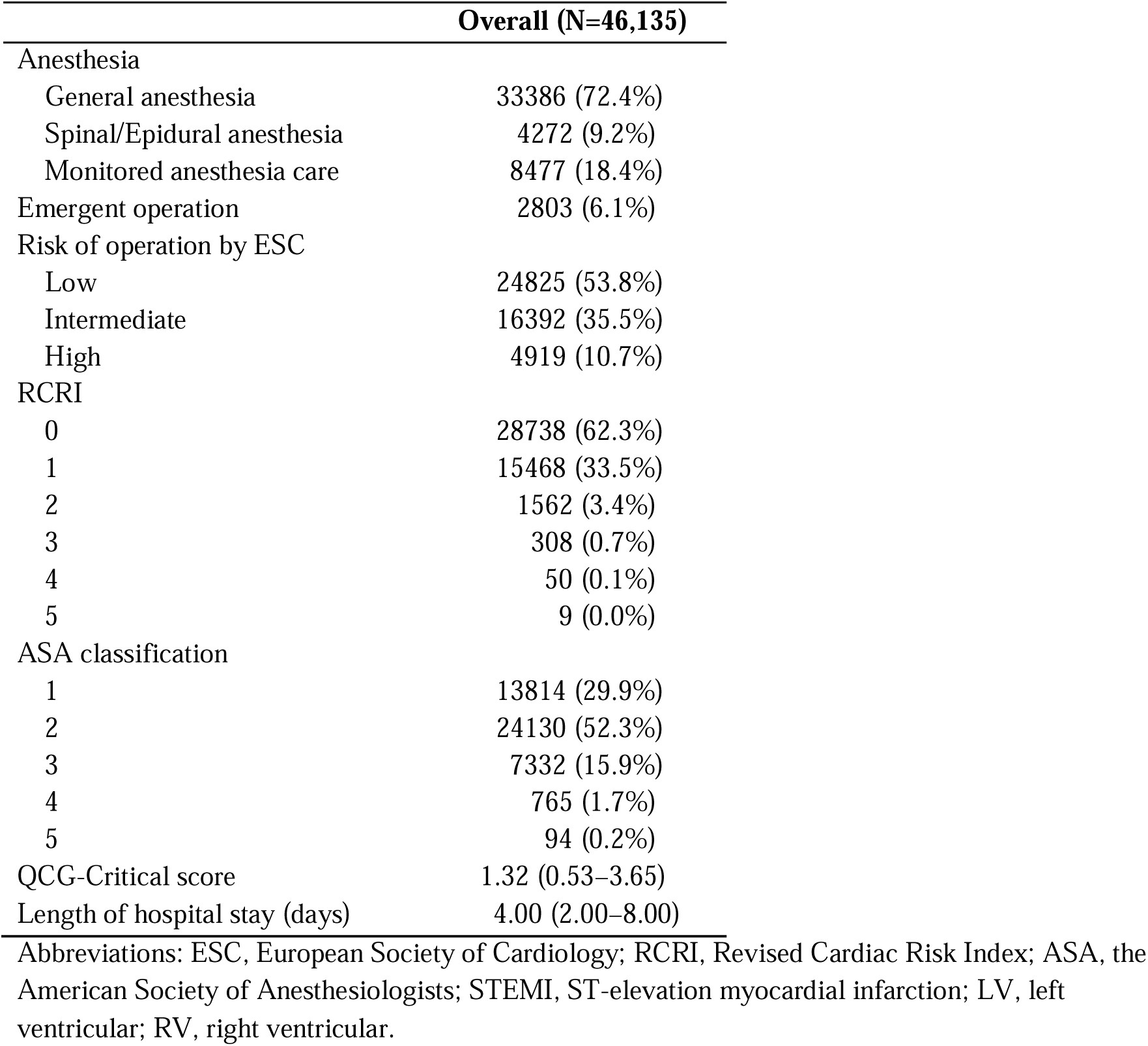
Surgery-specific baseline characteristics.

### Prevalence of postoperative events

Among the 46,135 surgeries, 156 in-hospital deaths occurred within 30 days of the index surgery, which corresponded to a mortality rate of 0.34%. Comparisons of demographics and surgery-specific factors between patients who survived beyond 30 days are presented in Supplementary Tables S2 and S3. Compared to the patients who survived beyond 30 days, participants in the mortality group were older, more likely to be male, and had a higher prevalence of comorbidities and abnormal laboratory findings than those in the control group. Moreover, participants in the mortality group showed higher surgery-specific parameters, including risk stratification according to ESC guideline as well as RCRI and ASA classification. The 30-day mortality rates of all surgeries and elective surgeries stratified by the operated body part are shown in Supplementary Figure S3.

### Establishment of the QCG-Critical score-based prediction model

The QCG-Critical scores that were derived from the ECG of the study population are presented in Table 2; the median (IQR) QCG-Critical score was 1.32 (0.53–3.65), and 4,326 cases (10.0%) had a QCG-Critical score >10 (Supplementary Table S4).

Univariable logistic regression showed that a higher QCG-Critical score indicated a significantly increased risk of postoperative mortality (OR 1.08 [1.07–1.09]; Supplementary Table S4). The analysis in the five subgroups stratified by the QCG-Critical score intervals (0–10, 10–20, 20–30, 30–40, and >40) showed a showed a dose-dependent increasing trend in the mortality rate. The subgroup with a QCG-Critical score >40 showed the highest mortality rate (11.7% vs. 0.1% in the subgroup with a QCG-Critical score <10; Figure 1).

**Figure 1.**
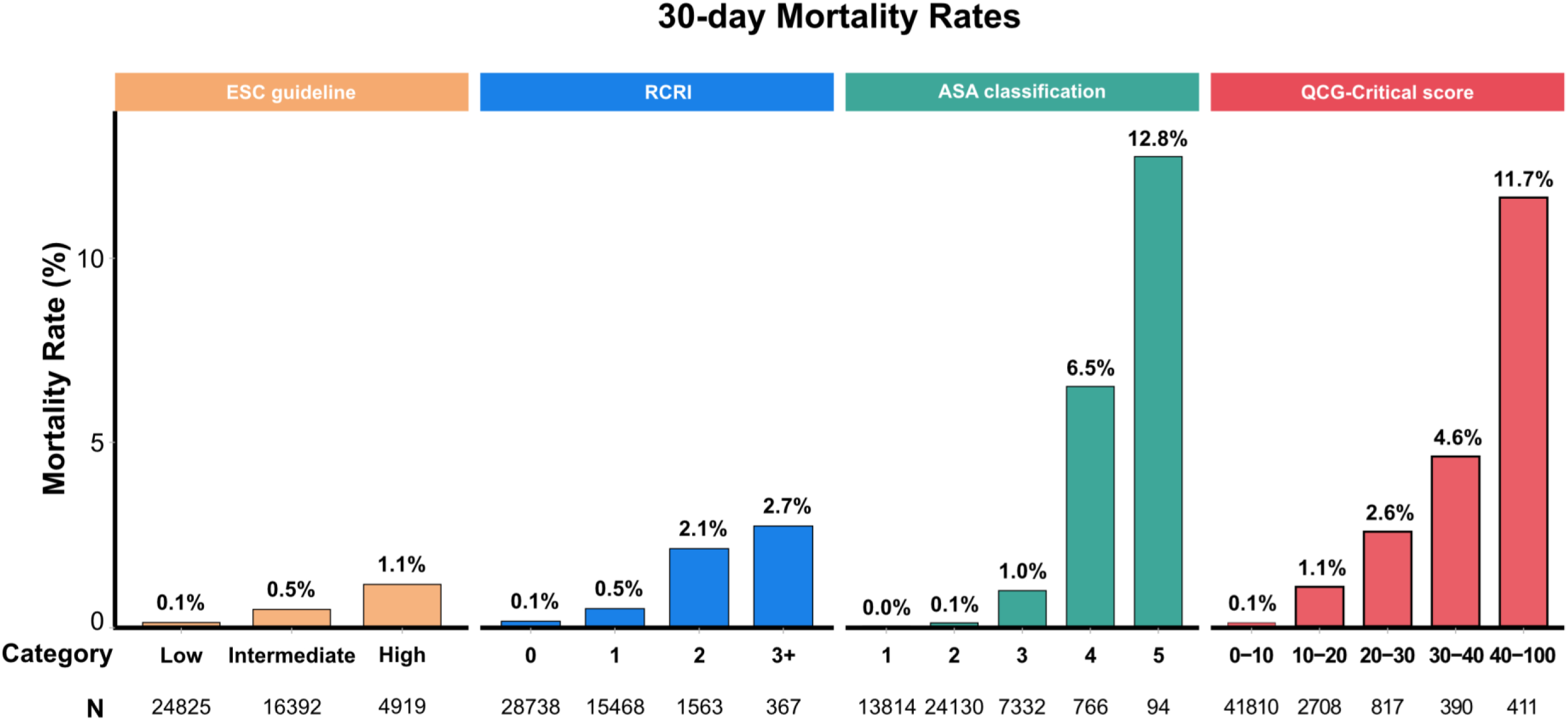
Comparison of 30-day postoperative mortality rates across different risk- stratification systems. The bar graphs show the 30-day in-hospital mortality rates according to the 2022 ESC guideline risk categories, RCRI scores, ASA physical status classification, and QCG-Critical score groups. Each stratification method showed an increasing trend in mortality in higher risk categories.

Furthermore, the prediction model that used the QCG-Critical score exhibited excellent performance, with sensitivity and specificity of 0.89 and 0.79, respectively, with an AUROC of 0.909 (95% CI). The accuracy of mortality prediction by QCG-Critical score remained consistent in the analysis that excluded 4,917 patients who underwent a second operation (AUROC 0.906, Supplementary Figure S4).

Next, we evaluated whether the performance of the QCG-based models improved after adding relevant clinical factors as covariates. Among clinical factors, older age (≥75 years), male sex, increased serum creatinine levels, and comorbidities were significantly associated with the primary outcome (Supplementary Table S4). However, the performance of the QCG-Critical score model did not significantly change after incorporating simple demographic factors and laboratory findings (Table 3). Therefore, for user convenience and simplicity, the final model included only the QCG-Critical score.

**Table 3.** Multivariable generalized estimating equation models using QCGcritical to predict the 30-day all-cause mortality.

|  | OR | 95% C.I. | p-value | AUROC |
| --- | --- | --- | --- | --- |
| **Model 1** |  |  |  | 0.909 |
| (Intercept) | 0.001 | 0.001–0.002 | <0.001 |  |
| QCG-Critical score | 1.08 | 1.07–1.08 | <0.001 |  |
| **Model 2** |  |  |  | 0.900 |
| (Intercept) | 0.001 | 0.001–0.001 | <0.001 |  |
| QCG-Critical score | 1.08 | 1.07–1.08 | <0.001 |  |
| Age ≥ 75 years | 2.25 | 1.56–3.23 | <0.001 | |
| Male sex | 2.20 | 1.51–3.22 | <0.001 |  |
| **Model 3** |  |  |  | 0.902 |
| (Intercept) | 0.001 | 0.001–0.001 | <0.001 |  |
| QCG-Critical score | 1.07 | 1.07–1.08 | <0.001 |  |
| Age ≥ 75 years | 1.99 | 1.39–2.86 | <0.001 | |
| Male sex | 2.01 | 1.38–2.94 | <0.001 |  |
| Creatinine ≥ 1.5 mg/dL | 2.07 | 0.52–1.73 | 0.001 | |
| Hemoglobin < 8 g/dL | 2.52 | 0.96–3.19 | 0.042 |  |
| Ischemic heart disease | 0.95 | 1.02–4.07 | 0.873 |  |
| Cerebrovascular accident | 1.75 | 0.67–3.51 | 0.067 |  |
| Heart failure | 2.03 | 1.33–3.22 | 0.045 |  |
| Diabetes mellitus using insulin | 1.53 | 1.04–6.15 | 0.310 |  |

### Predictive performance of the QCG-score model as compared to conventional models

We compared the performance of the prediction models using the QCG-Critical score and traditional perioperative risk-stratification tools. The novel perioperative risk-prediction model that utilized the QCG-Critical score significantly outperformed the models that used the ESC guideline or RCRI (AUROCs: QCG-Critical score, 0.909; ESC guideline, 0.728; and RCRI, 0.725; *p*<0.05 for comparisons with both the ESC guideline and RCRI); however, the performance of the prediction model that used the ASA classification was comparable to that of the novel QCG-Critical score-based model (AUROC for ASA classification: 0.886; *p* = 0.090; Figure 2). The QCG-Critical score model demonstrated excellent calibration with a bias-corrected curve that closely aligned with the ideal line and a mean absolute error of 0.002 (Supplementary Figure S5). The effect of the QCG-Critical score remained consistent in various subgroups stratified by age, sex, emergency operation, anaesthesia method, and risk groups that were identified based on conventional methods (Figure 3). Although statistically significant interactions were observed in selected subgroups, such as ASA class, ESC guidelines, and RCRI, the score remained predictive of mortality in all subgroups.

**Figure 2.**
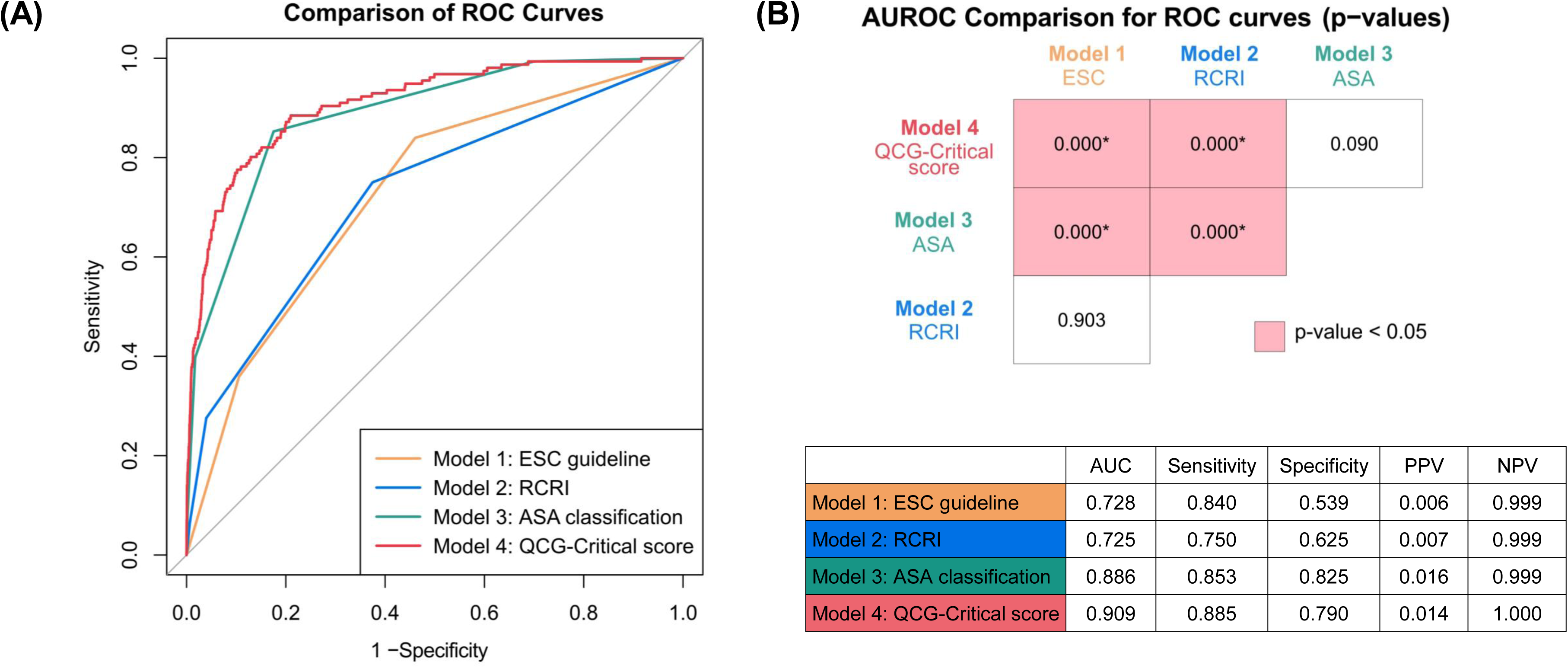
Comparison of receiver operating characteristic (ROC) curves and area under the ROC curve (AUROC) values among different perioperative risk models. (A) ROC curves for 30-day postoperative mortality prediction using the ESC guideline, RCRI, ASA class, and QCG-Critical score models. (B) Pairwise AUROC comparison matrix with *p*-values from DeLong’s test, indicating the statistical significance of the QCG-Critical score’s superiority over the ESC and RCRI models. The accompanying table shows the AUROC, Youden Index, sensitivity, specificity, positive predictive value (PPV), and negative predictive value (NPV) for each model.

**Figure 3.**
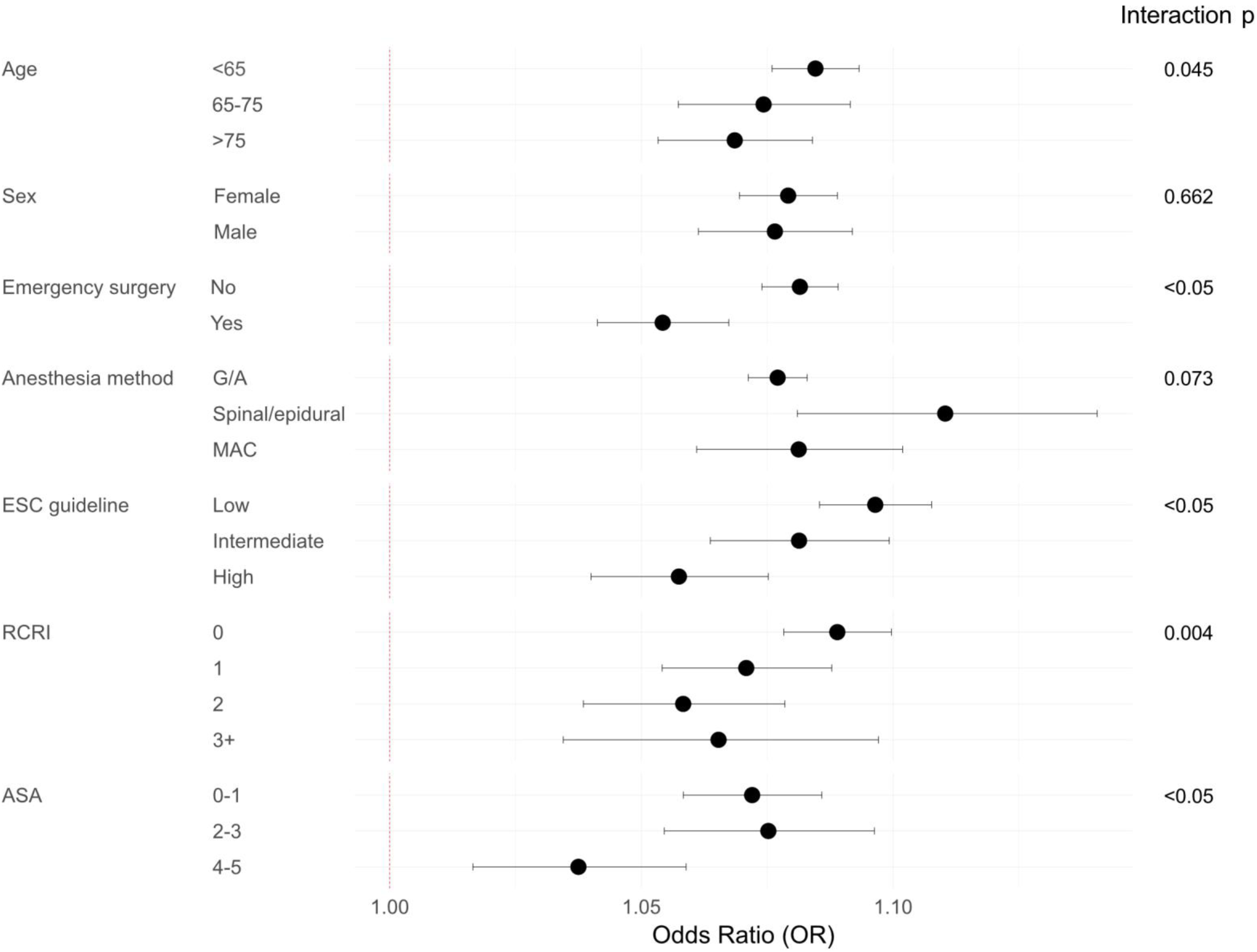
Subgroup analysis of the QCG-Critical score model for predicting 30-day mortality. The Forest plot shows odds ratios with 95% confidence intervals for the association between the QCG-Critical score and 30-day mortality across various subgroups, stratified by age, sex, emergency surgery, ESC risk category, anaesthesia method, RCRI, and ASA classification. Interaction *p*-values are displayed for heterogeneity across subgroups. The QCG-Critical score remained significantly predictive in all groups and demonstrated minimal variation in performance.

### Secondary outcomes

Beyond 30-day mortality, the QCG-Critical score demonstrated a strong predictive performance for secondary outcomes (Figure 4). Notably, the AUROC for 7-day mortality was 0.933, which exceeded the performance of the model for the primary outcome and indicated the superior short-term prognostic value of the QCG-Critical score.

**Figure 4.**
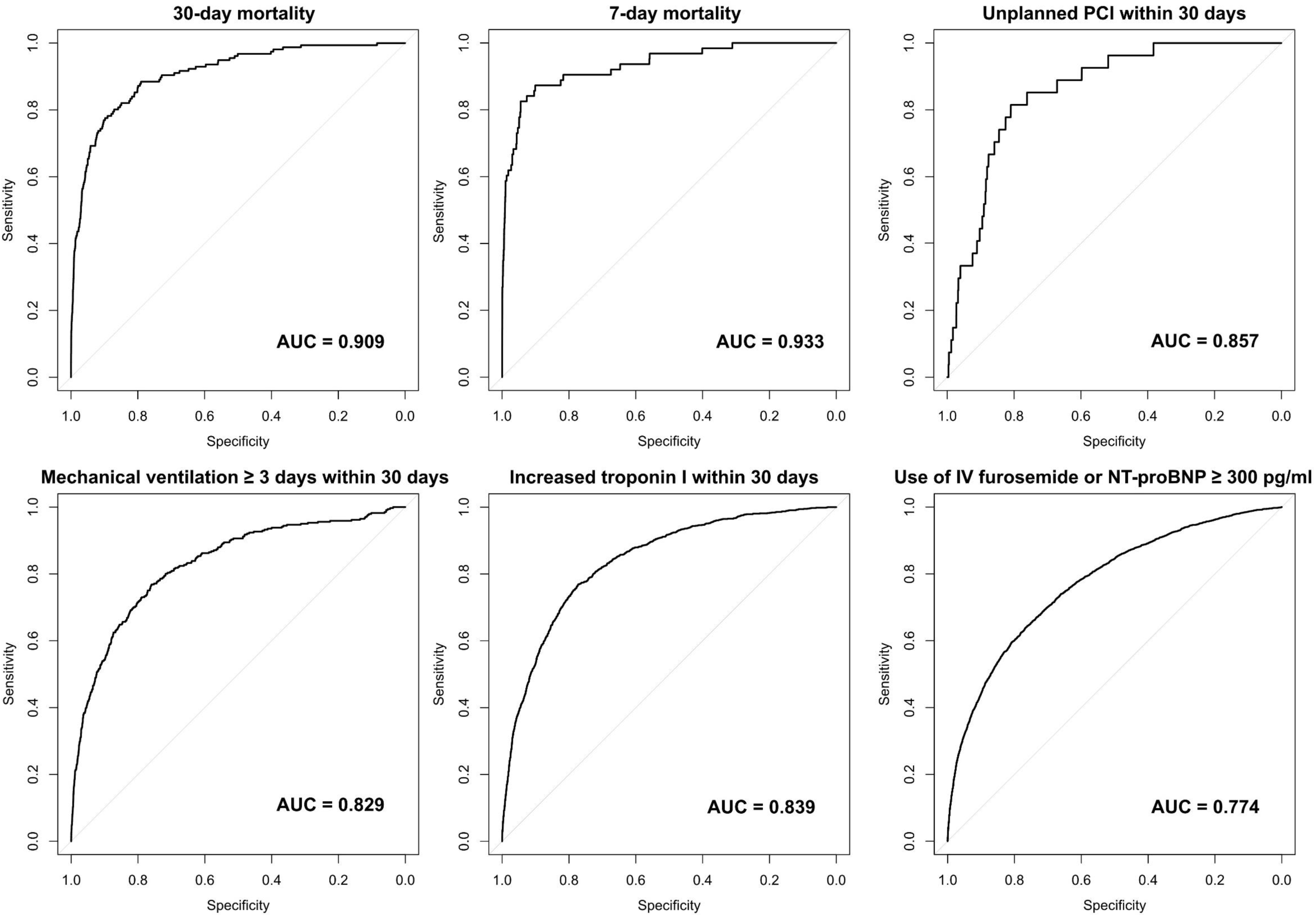
ROC curves for the prediction of secondary postoperative outcomes using the QCG-Critical score. (A) 30-day mortality, (B) 7-day mortality, (C) unplanned percutaneous coronary intervention (PCI) within 30 days, (D) mechanical ventilation ≥3 days, (E) increased troponin I within 30 days, and (F) use of intravenous (IV) furosemide or N-terminal pro B-type natriuretic peptide (NT-proBNP) ≥300 pg/mL.

## DISCUSSION

In this study, we evaluated the utility of an AI-derived ECG parameter, the QCG-Critical score, for predicting postoperative 30-day in-hospital mortality and compared its performance with that of traditional perioperative risk-assessment tools. Unlike existing models that require several clinical and laboratory data points, the QCG-Critical score demonstrated a high predictive performance while using only ECG data and outperformed the risk-stratification systems that were based on the ESC guideline and RCRI (Supplementary Table S1).

Recent guidelines on preoperative risk assessment recommend the use of a 12-lead ECG for patients undergoing intermediate- or high-risk non-cardiac surgery, particularly in those who are suspected of having cardiovascular diseases or patients who have cardiovascular risk factors.^2,3^ Nevertheless, the low cost and non-invasive nature of ECG allow its widespread use in preoperative evaluations, regardless of individual risk.^7^ However, the quantity and quality of information obtained from ECG are highly dependent on the physician’s experience and the patient’s clinical features.^17,18^ The indiscrete use of ECG as a preoperative examination could lead to unnecessary additional tests, delays in surgical planning, and resource wastage.^19,20^

In this study, however, the QCG-Critical score that was derived using an AI-ECG analysis tool showed excellent predictive performance in detecting patients with a high risk of postoperative mortality, and performed significantly better than conventional risk-stratification systems. For example, the ESC guideline classified more than 10% of the study population into the ‘high-risk’ group whereas the prevalence of postoperative mortality in this subgroup did not significantly differ from that in the lower-risk groups. In contrast, based on the QCG-Critical score, the highest-risk group (>40) comprised only 1% of the population whereas there was a 11.7% risk for the primary outcome. This result suggests that the novel AI-based system enables the efficient selection of patients who require a more intensive preoperative evaluation, which ultimately leads to a significant reduction in the unnecessary use of medical resources. The comparable performance of the QCG-Critical score to the ASA system is encouraging; however, several reports have pointed out the inter-rater variability of the ASA classification system, which especially depends on the anaesthesiologist’s experience.^21–23^ The QCG-Critical score can provide objective risk assessment independently of the evaluator, which supports its potential benefits in real-world practice. Furthermore, the QCG-Critical score demonstrated a consistent predictive performance across a wide range of clinical subgroups. Although the magnitude of the association was attenuated in patients undergoing emergency surgery or those classified by conventional tools as having higher risk, the score remained significantly associated with postoperative mortality in all subgroups.

Notably, the addition of relevant clinical factors to the single QCG-Critical score-based model did not significantly change the performance. Previous studies on AI-ECG models have shown that AI-enhanced ECG can predict various clinical features of patients, such as age, sex, body mass index, hyperkalaemia, echocardiographic findings, and even the risk of cardiac diseases.^8^ Thus, the ECG can be compared to a one-page encrypted summary of a person’s current health status that assimilates various aspects, such as demographic factors, comorbidities, and laboratory findings, of the individual’s status at a single time point.

Despite the limitations of decoding this image through the human eye, AI-based decoding can unpack hidden information to enable greater information recovery. Though developing a more complex model may slightly improve predictive accuracy, implementing a highly complex risk-stratification tool may not be practical in real-world clinical settings.

Several studies have used AI and ML algorithms to predict adverse postoperative outcomes. ^24–26^ Despite the excellent performance of these models in predicting postoperative mortality, most used various clinical parameters extracted from electrical healthcare record (EHR) database. Therefore, they require extensive input to analyse the probability, and their performance inevitably relies on the quality of data regarding subjective impressions (e.g., functional classification) in medical records. In contrast, the QCG model requires only an ECG image file or even a photograph obtained using a smartphone,^15^ which ensures the ease of applicability of the model in resource-limited clinical settings. Furthermore, the QCG- Critical score can be generated in less than one second and is displayed alongside ECG images, in a similar way to automated ECG interpretations that are generated by ECG machines. Therefore, the QCG system enables rapid and objective patient-specific perioperative risk assessment within EHR systems.

A recent study using an AI-enhanced ECG model demonstrated its excellent performance in predicting postoperative outcomes.^27^ Though this model was specifically trained for postoperative mortality, the QCG-Critical score was originally developed to identify critically ill patients in the emergency department. Nevertheless, our model demonstrated strong performance in perioperative risk assessment without additional modifications as well as in predicting other important postoperative outcomes, such as 7-day mortality, unplanned PCI, mechanical ventilation, increased troponin I, and presumed heart failure. We previously demonstrated that the QCG-Critical score could predict in-hospital cardiac mortality in patients admitted for acute decompensated heart failure.^13^ This suggests that a general AI-enhanced ECG model, which is not trained for specific outcomes, can be applied to predict diverse postoperative outcomes without incorporating major modifications to clinical variables.

Despite its strengths, some limitations of the study warrant mention. Though the data collection period of this study did not overlap with the training data collection period, it is not uncommon for the accuracy of models to be reduced in different populations, and this emphasizes the importance of external validation of models. Considering the single-centre retrospective study design, potential concerns regarding unmeasured bias may arise owing to the composition of surgery types and variations in clinical practice among individual surgeons and anaesthesiologists. Therefore, external validation in diverse clinical settings is required to confirm the generalizability of the findings. In addition, our study population showed a significantly lower postoperative mortality rate, which could have influenced the performance of the model. As the overall surgical risk has been gradually declining owing to advancements in surgical techniques, enhanced surveillance systems, and improved postoperative care, despite the increase in the population with high comorbidities,^4,5^ our QCG-Critical score model potentially constitutes a better short-term prognosticator in the low-postoperative mortality era. Finally, though we demonstrated the consistent accuracy of the model in various subgroups, our results should be replicated in future large-scale studies. Moreover, because some types of surgery involve a small number of patients and exhibit very low mortality rates, the generalization of the results could be premature.

In conclusion, an AI-derived QCG-Critical score derived from ECG demonstrated high predictability for 30-day postoperative mortality and other adverse outcomes, and outperformed pre-existing perioperative risk-stratification tools. With its high accuracy, objectivity, and ease of integration into routine preoperative workflows, the QCG-Critical scoring system has the potential to enhance perioperative risk stratification without additional clinical inputs.

### Clinical Perspectives

#### Competency in Medical Knowledge

An AI-derived ECG risk score (QCG-Critical score) can accurately predict postoperative mortality and complications using only preoperative ECG images, offering a novel non-invasive tool for perioperative risk stratification in non-cardiac surgery.

#### Translational Outlook 1

The QCG-Critical score may be integrated into routine surgical workflows as an automated, objective, and rapid triage tool, especially in resource-limited settings where traditional risk scores may be impractical.

#### Translational Outlook 2

Future multicenter prospective studies are needed to externally validate the QCG-Critical score and determine its clinical impact on surgical planning, monitoring intensity, and postoperative outcomes.

## Data Availability

The data used in the present study cannot be made publicly available owing to ethical restrictions set by the Seoul National University Bundang Hospital Institutional Review Board (https://e-irb.snubh.org), as patient and participant privacy would be compromised. A minimally anonymized dataset can be requested by contacting the corresponding authors.

## Funding

This research was supported by a grant from the Korea Health Technology R&D Project through the Korea Health Industry Development Institute (KHIDI), which is funded by the Ministry of Health & Welfare, Republic of Korea (grant number: RS-2023-00265933).

## Disclosures

Joonghee Kim developed the algorithm and is the founder and CEO of the startup company, ARPI Inc. Youngjin Cho has worked at ARPI Inc. as a research director. The remaining authors declare no conflicts of interest.

## Acknowledgment

None

## Abbreviations

AI: artificial intelligence
ASA: the American Society of Anesthesiologists
AUROC: area under the receiver operating characteristic curve
ECG: Electrocardiography
EHR: electrical healthcare record
ESC: European Society of Cardiology
ML: machine learning
PCI: percutaneous coronary intervention
RCRI: Revised Cardiac Risk Index

## Author Contributions

All authors were involved in the conceptualization and design of this research. YK and JP were involved in data collection, and H-MC, YK, JK, JHL, and I-YO analyzed the data. YEY, I-YO, and JHL were responsible for investigation. JK and YC were responsible for the generation of the QCG score. H-MC and YK wrote the manuscript draft and prepared figures, and YEY, JK, YC, and I-AS reviewed and corrected the manuscript. All authors approved the final version of the manuscript.

## Central Illustration. AI-ECG for Perioperative Risk Prediction

This study evaluated the QCG-Critical score, an AI-derived risk score generated from preoperative ECG images, in 46,135 non-cardiac surgeries. The 30-day postoperative mortality risk increased with higher QCG-Critical scores. The QCG-Critical score showed strong predictive performance for 30-day mortality and other adverse events, including unplanned percutaneous coronary intervention, mechanical ventilation, troponin elevation, and heart failure. The QCG model outperformed conventional risk tools such as the ESC guideline, RCRI, and ASA classification, with an AUROC of 0.909.

Abbreviations: ASA, the American Society of Anesthesiologists; AUROC, area under the receiver operating characteristic curve; ESC, European Society of Cardiology; RCRI, Revised Cardiac Risk Index.

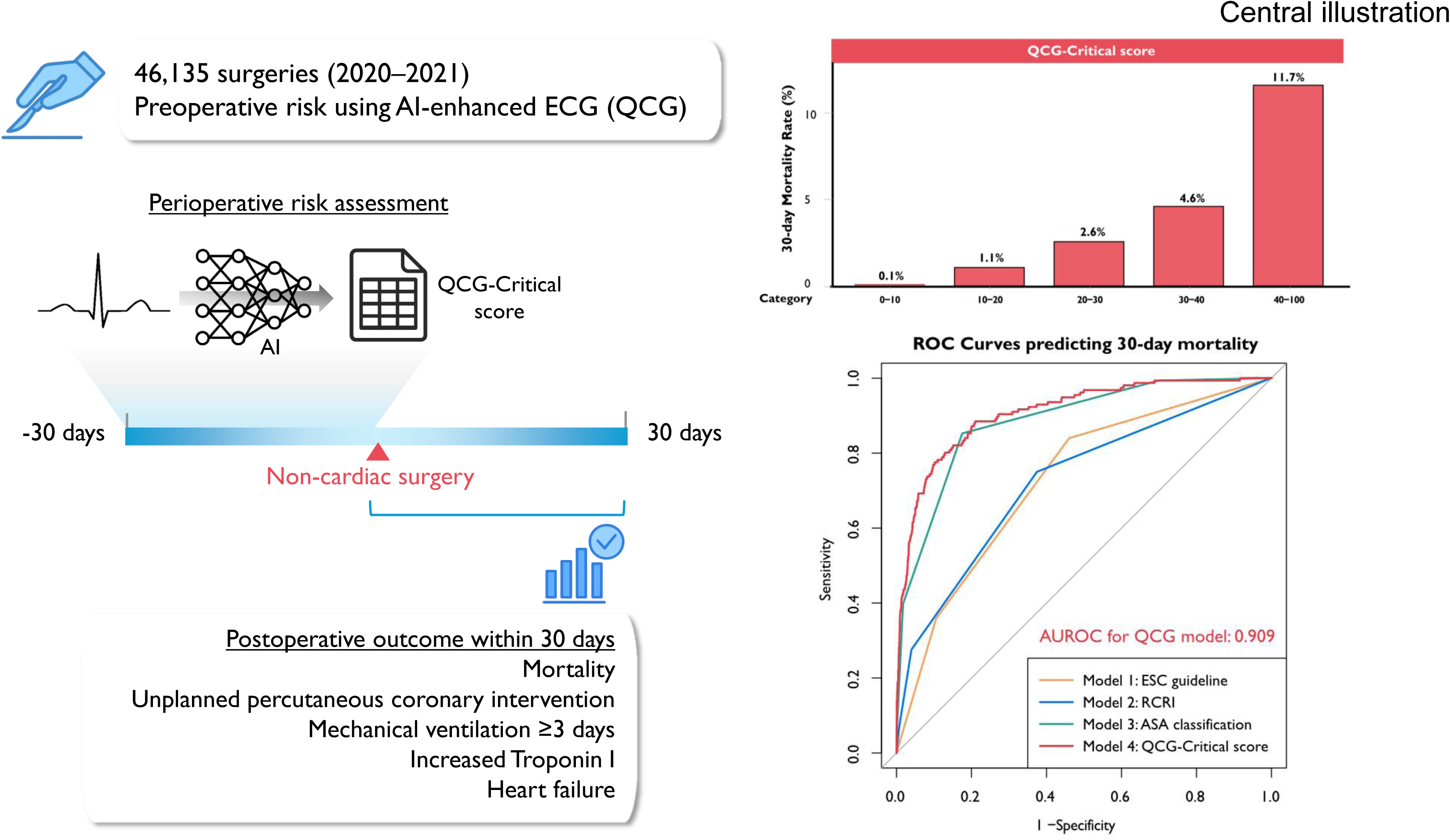

## Notes

### Author Declarations

The study protocol was approved by the Institutional Review Board of Seoul National University Bundang Hospital (IRB No. B-2409-927-106), which waived the requirement for written informed consent owing to the retrospective nature of the study.

