## Supplementary file for "Artificial intelligence-enhanced Electrocardiography Score for Perioperative Risk Assessment in Non-cardiac Surgery"

**Supplementary Table S1. Comparison between traditional perioperative risk stratification tools and QCG-Critical score**

| Perioperative risk stratification tools | Operative risk by European Society of Cardiology (2022)^[1]^ | Revised Cardiac Risk Index (RCRI)^[2]^ | American Society of Anesthesiologists Physical Status Class^[3]^ | QCG-Critical score |
| --- | --- | --- | --- | --- |
| Criteria | Low / Intermediate / High risk according to the surgery-related risk | - High-risk surgery (intrathoracic, intra-abdominal, suprainguinal vascular surgery)  - Ischemic heart disease  - Congestive heart failure  - Cerebrovascular disease  - Renal Insufficiency (Cr > 2mg/dl)  - Diabetes treated with insulin | I = A normal healthy patient  II = A patient with mild systemic disease  III = A patient with severe systemic disease  IV = A patient with severe systemic disease that is a constant threat to life  V = A moribund patient who is not expected to survive without the operation | QCG-Critical score from AI-enhanced ECG |
| Methods | Categorize based on the type of surgery | Sum of the points  (1 point per risk factor) | Subjective assessment by anesthesiologists |  |
| Outcome | Broad approximation of 30-day risk of CV death, MI, and stroke, which only considers the specific surgical intervention without  considering the patient’s comorbidities | MI/Cardiac Arrest, complete heart block, pulmonary edema during admission |  | 30-day mortality |
| Predicted probability of outcomes | Low risk: < 1%  Intermediate risk: 1-5%  High risk: > 5% | 1: risk 6.0% (4.9–7.4)  2: risk 10.1% (8.1–10.6)  ≥3: risk 15% (11.1–20.0) |  | 25: 0.97%  30: 1.41%  40: 2.98%  50: 6.18%  60: 12.39% |

**Supplementary Table S2. Baseline characteristics of patients according to survival status**

|  | **Alive (N=45,979)** | **Death  (N=156)** | **P-value** |
| --- | --- | --- | --- |
| Age | 56.51 (16.13) | 69.33 (12.18) | <0.001 |
| Male Sex | 20449 (44.5) | 112 (71.8) | <0.001 |
| Previous history |  |  |  |
| Diabetes mellitus | 5868 (12.8) | 36 (23.1) | <0.001 |
| Hypertension | 10704 (23.3) | 55 (35.3) | 0.001 |
| Heart failure | 399 (0.9) | 11 (7.1) | <0.001 |
| Ischemic heart disease | 1753 (3.8) | 19 (12.2) | <0.001 |
| Cerebrovascular event | 1505 (3.3) | 20 (12.8) | <0.001 |
| Smoking |  |  |  |
| Nonsmoker | 28332 (61.6) | 104 (66.7) | <0.001 |
| Smoker | 9852 (21.4) | 52 (33.3) |  |
| Unknown | 7795 (17.0) | 0 (0.0) |  |
| Laboratory findings |  |  |  |
| Hemoglobin (g/dL) | 13.27 (1.86) | 11.34 (2.26) | <0.001 |
| WBC count (×10³/μL) | 6.44 (5.33–7.85) | 9.38 (6.13–13.12) | <0.001 |
| Platelet count (×10³/μL) | 241 (201–286) | 192 (121–265) | <0.001 |
| BUN (mg/dL) | 15.00 (12.00–18.00) | 20.00 (14.00–33.25) | <0.001 |
| Creatinine (mg/dL) | 0.74 (0.61–0.91) | 0.89 (0.67–1.67) | <0.001 |
| AST (U/L) | 25.00 (20.00–31.00) | 33.00 (24.00–56.25) | <0.001 |
| ALT (U/L) | 20.00 (14.00–30.00) | 21.00 (13.00–40.50) | 0.263 |
| Albumin (g/dL) | 4.30 (4.10–4.50) | 3.60 (2.90–4.10) | <0.001 |
| PT INR | 0.96 (0.93–1.01) | 1.12 (1.02–1.38) | <0.001 |

Abbreviations: WBC, white blood cell; BUN, blood urea nitrogen; AST, aspartate aminotransferase; ALT, Alanine transaminase; PT INR, prothrombin time/international normalized ratio.

**Supplementary Table S3. Surgery-specific baseline characteristics according to survival status**

|  | **Alive (N=45,979)** | **Death  (N=156)** | **P-value** |
| --- | --- | --- | --- |
| Anesthesia |  |  | <0.001 |
| General anesthesia | 33244 (72.3) | 142 (91.0) |  |
| Spinal/Epidural anesthesia | 4268 (9.3) | 4 (2.6) |  |
| Monitored anesthesia care | 8467 (18.4) | 10 (6.4) |  |
| Emergent operation | 2728 (5.9) | 75 (48.1) | <0.001 |
| Risk of operation by ESC |  |  |  |
| Low | 24800 (53.9) | 25 (16.0) | <0.001 |
| Intermediate | 16317 (35.5) | 75 (48.1) |  |
| High | 4862 (10.6) | 56 (35.9) |  |
| RCRI |  |  |  |
| 0 | 28699 (62.4) | 39 (25.0) | <0.001 |
| 1 | 15394 (33.5) | 74 (47.4) |  |
| 2 | 1529 (3.3) | 33 (21.2) |  |
| 3 | 298 (0.6) | 10 (6.4) |  |
| 4 | 50 (0.1) | 0 (0.0) |  |
| 5 | 9 (0.0) | 0 (0.0) |  |
| ASA classification |  |  | <0.001 |
| 1 | 13813 (30.0) | 1 (0.6) |  |
| 2 | 24108 (52.4) | 22 (14.1) |  |
| 3 | 7261 (15.8) | 71 (45.5) |  |
| 4 | 715 (1.6) | 50 (32.1) |  |
| 5 | 82 (0.2) | 12 (7.7) |  |
| QCG-Critical score | 1.31 (0.53–3.61) | 21.49 (9.82–46.23) | <0.001 |
| Length of hospital stay (days) | 6.55 (13.71) | 16.65 (13.79) | <0.001 |

Abbreviations: ESC, European Society of Cardiology; RCRI, Revised Cardiac Risk Index; ASA, the American Society of Anesthesiologists; STEMI, ST-elevation myocardial infarction; LV, left ventricular; RV, right ventricular

**Supplementary Table S4. Univariable generalized estimating equation models to predict the 30-day all-cause mortality**

| **Variable** | **OR** | **95% C.I.** | **P-Value** |
| --- | --- | --- | --- |
| Age | 1.06 | 1.05–1.08 | <0.001 |
| Age ≥ 65 years | 3.52 | 2.53–4.91 | <0.001 |
| Age ≥75 years | 4.09 | 2.96–5.65 | <0.001 |
| Male sex | 3.20 | 2.26–4.54 | <0.001 |
| Previous history |  |  |  |
| Diabetes mellitus | 2.06 | 1.42–2.99 | <0.001 |
| Diabetes mellitus using insulin | 5.01 | 2.63–9.54 | <0.001 |
| Hypertension | 1.81 | 1.3–2.52 | <0.001 |
| Heart failure | 8.67 | 4.68–16.07 | <0.001 |
| Ischemic heart disease | 3.55 | 2.19–5.75 | <0.001 |
| Cerebrovascular event | 4.43 | 2.76–7.12 | <0.001 |
| Laboratory findings |  |  |  |
| Hemoglobin | 0.63 | 0.59–0.68 | <0.001 |
| Hemoglobin < 8 g/dL | 8.42 | 3.79–18.72 | <0.001 |
| Hemoglobin < 10 g/dL | 7.93 | 5.63–11.17 | <0.001 |
| Hemoglobin < 11 g/dL | 6.46 | 4.69–8.88 | <0.001 |
| Hemoglobin < 12 g/dL | 4.90 | 3.56–6.73 | <0.001 |
| WBC | 1.16 | 1.13–1.19 | <0.001 |
| Platelet | 0.99 | 0.99–1.00 | <0.001 |
| BUN | 1.04 | 1.03–1.04 | <0.001 |
| Creatinine | 1.23 | 1.18–1.28 | <0.001 |
| Creatinine ≥ 1.5 mg/dL | 8.35 | 5.81–12.01 | <0.001 |
| Creatinine ≥ 2.0 mg/dL | 7.70 | 5.16–11.5 | <0.001 |
| AST | 1.01 | 1.00–1.01 | <0.001 |
| ALT | 1.00 | 1.00–1.00 | <0.001 |
| PT INR | 1.03 | 1.03–1.04 | <0.001 |
| Anesthesia |  |  |  |
| General anesthesia | 3.91 | 2.25–6.78 | <0.001 |
| Spinal or epidural anesthesia | 0.26 | 0.09–0.7 | 0.008 |
| MAC | 0.30 | 0.16–0.58 | <0.001 |
| Emergent surgery | 14.75 | 10.77–20.19 | <0.001 |
| Risk of surgery by ESC |  |  |  |
| Low risk by ESC 2022 guideline | 0.17 | 0.11–0.25 | <0.001 |
| Intermediate risk by ESC 2022 guideline | 1.67 | 1.22–2.3 | 0.001 |
| High risk by ESC 2022 guideline | 4.72 | 3.4–6.55 | <0.001 |
| RCRI |  |  |  |
| RCRI ≥ 2 | 4.96 | 3.45–7.14 | <0.001 |
| RCRI ≥ 3 | 8.92 | 6.27–12.7 | <0.001 |
| ASA classification | 1.86 | 5.57–7.43 | <0.001 |
| ASA ≥ 3 | 27.22 | 17.48–42.41 | <0.001 |
| QCG-Critical score | 1.08 | 1.07–1.09 | <0.001 |
| QCG-Critical score ≥ 1 | 21.91 | 9.07–52.96 | <0.001 |
| QCG-Critical score ≥ 2.5 | 22.03 | 12.5–38.82 | <0.001 |
| QCG-Critical score ≥ 5 | 22.56 | 14.7–34.63 | <0.001 |
| QCG-Critical score ≥ 10 | 28.72 | 20.01–41.23 | <0.001 |
| QCG-Critical score ≥ 20 | 36.55 | 26.5–50.41 | <0.001 |

Abbreviations are consistent with those in Supplementary Table S1 and S2.

**Supplementary Figure 1. Study flow**

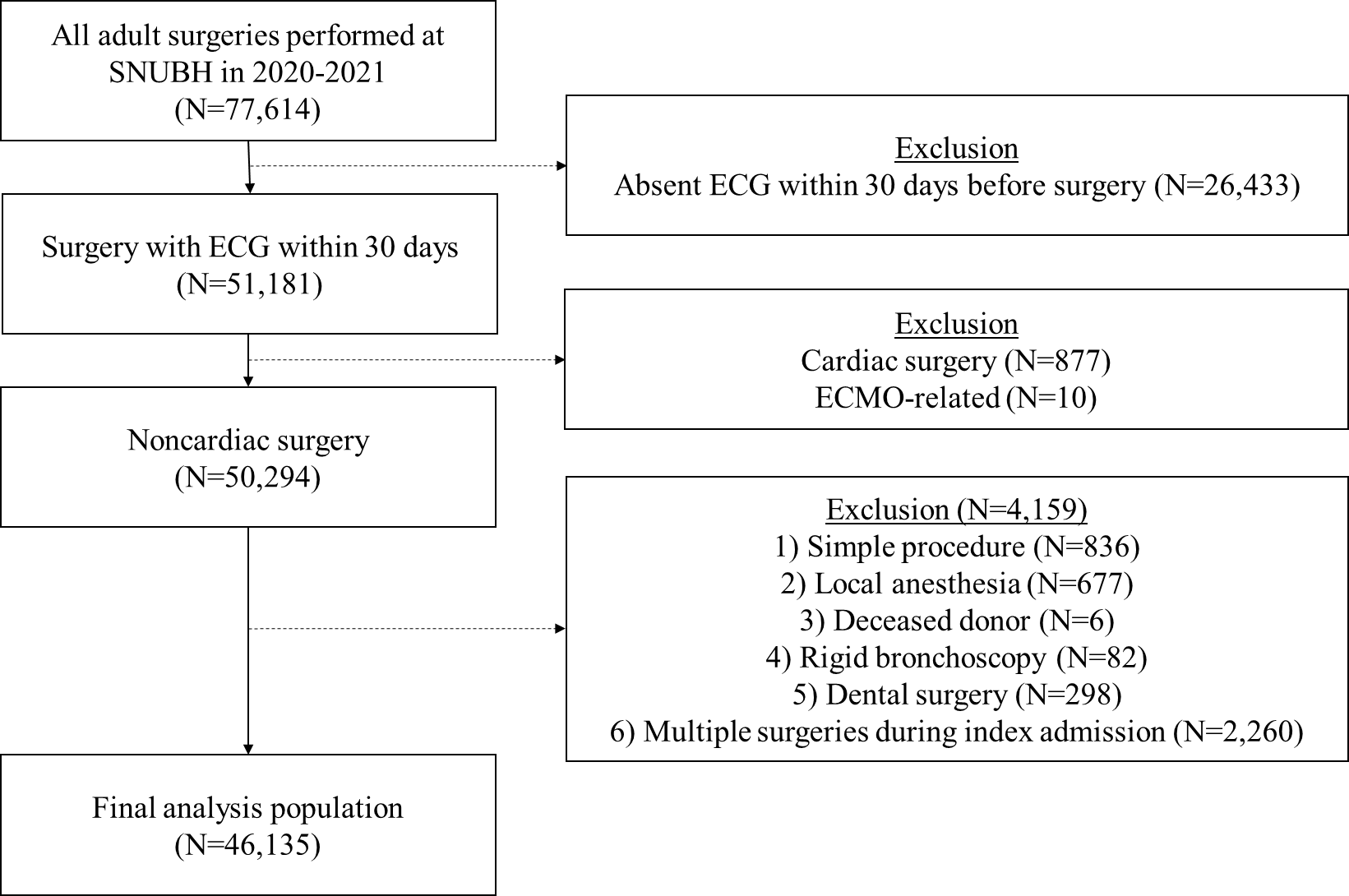

**Supplementary Figure 2. Classification of surgeries and number of patients included in the study according to the department**

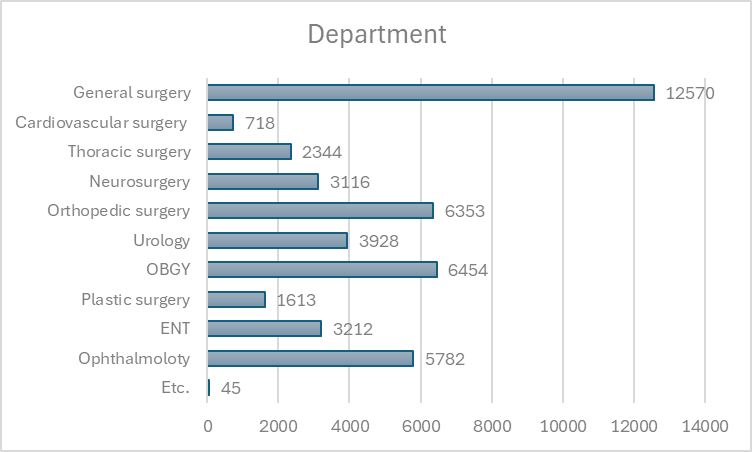

**Supplementary Figure 3. The number of patients and 30-day mortality by body part**

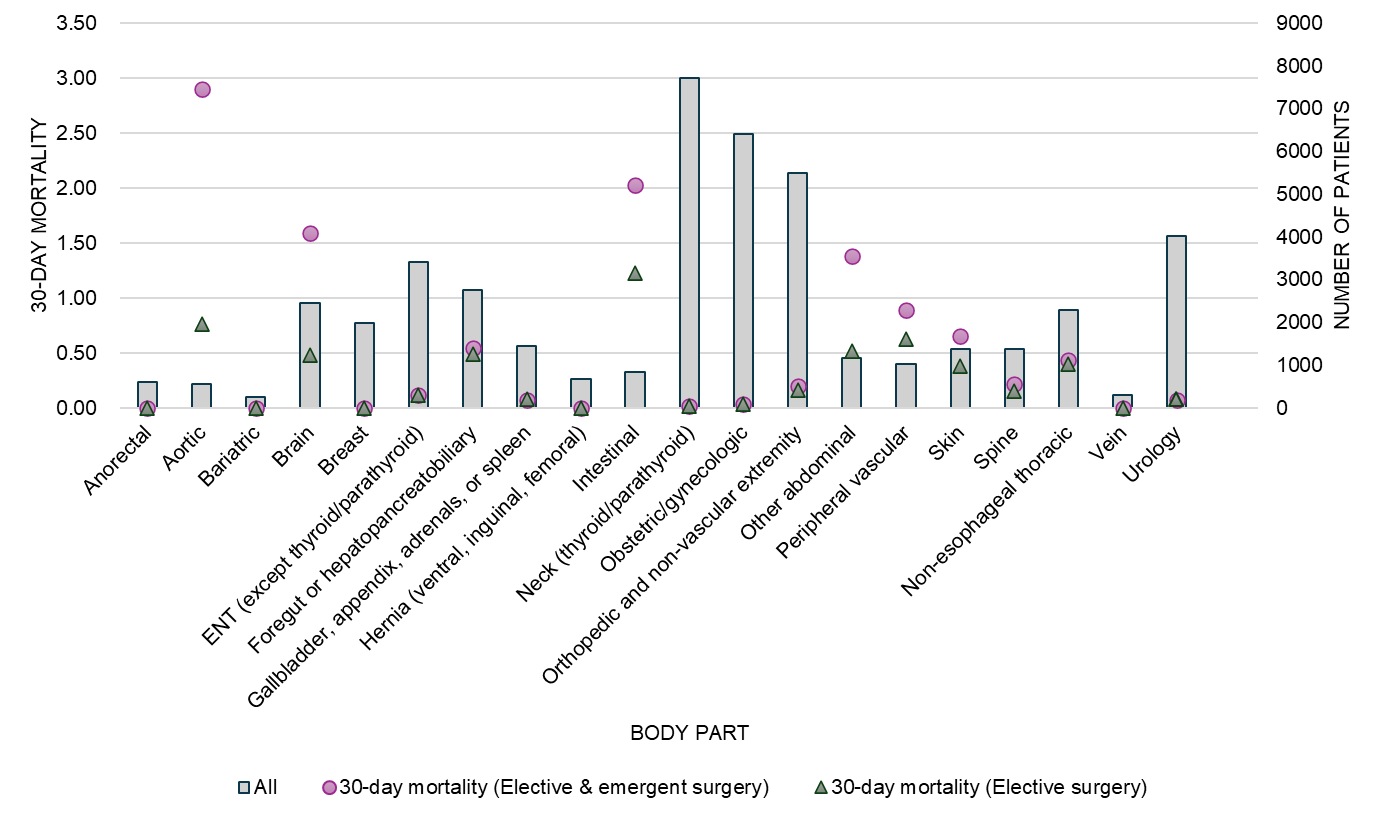

**Supplementary Figure 4. Sensitivity analysis for 41,218 patients after exclusion of multiple surgeries**

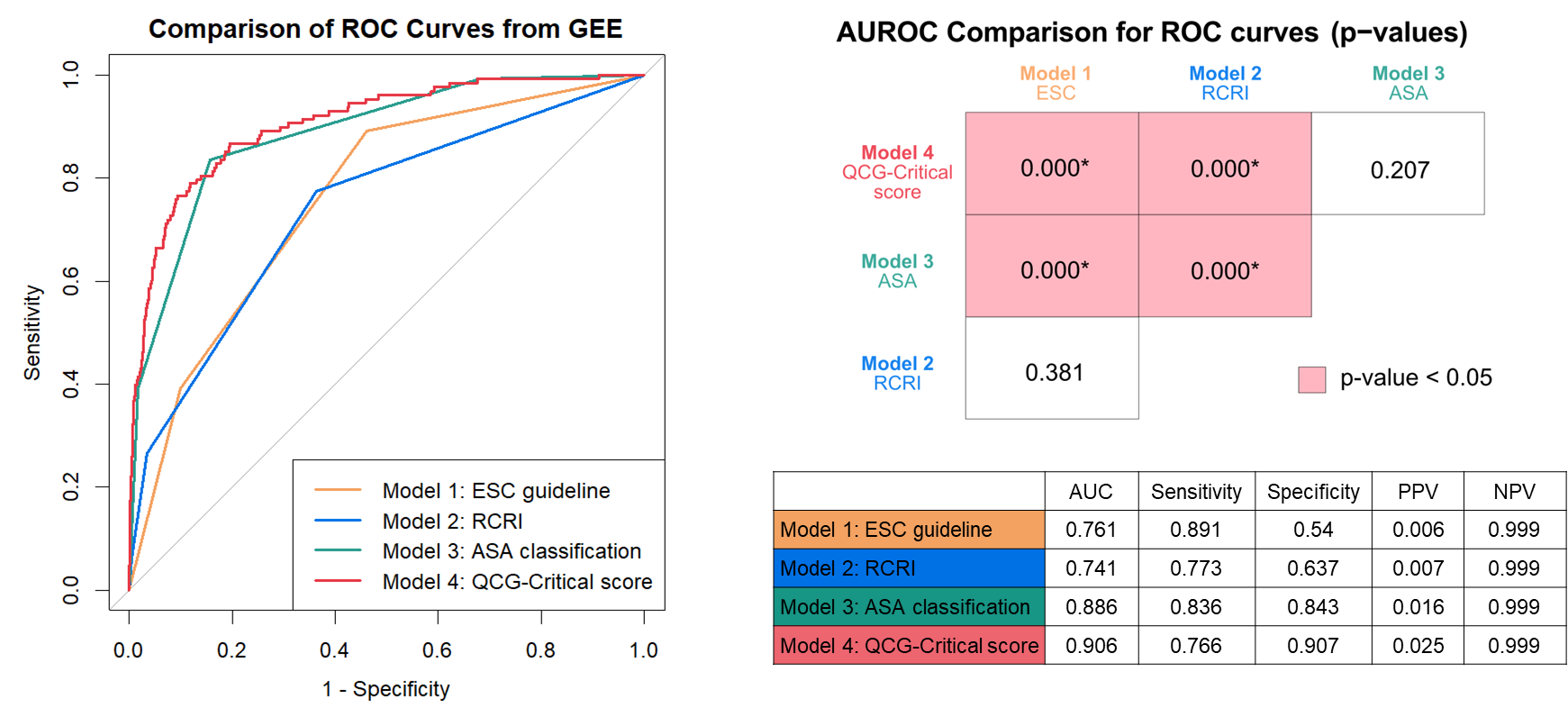

**Supplementary Figure 5. Calibration plot of QCG-Critical score model**

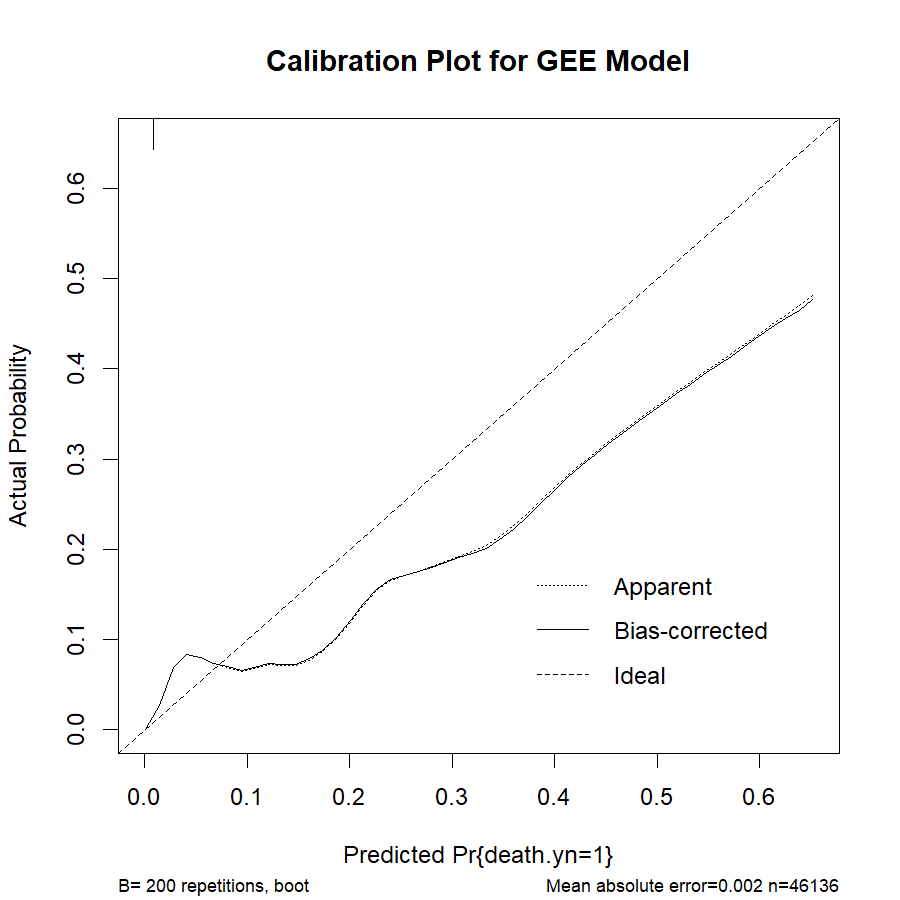

1. Halvorsen S, Mehilli J, Cassese S, Hall TS, Abdelhamid M, Barbato E. 2022 ESC Guidelines on cardiovascular assessment and management of patients undergoing non-cardiac surgery. Eur Heart J. 2022;43(39):3826–924.
2. Derivation and Prospective Validation of a Simple Index for Prediction of Cardiac Risk of Major Noncardiac Surgery. Circulation. 1999;100:1043–9.
3. A review of ASA physical status - historical perspectives and modern developments. Anaesthesia. 2019 Mar 15;74(3):373–9.
